# The Data Burden of Digital Learning

**DOI:** 10.1101/2024.08.23.24312503

**Authors:** Rachel Umoren, Ime Asangansi, Dillon Afenir, Brian W. Bresnahan, Annabelle Kotler, Cailin White, Matt Cook, Casey Lowman, Sara Berkelhamer

## Abstract

The costs of participating in training programs that rely on video conferencing vary by mechanics of use and the specific platform. We proposed practical solutions to limiting costs in low resource settings with the use of video conferencing calls. Scenarios in which facilitators have their video on and expect learners to participate with continuous video result in the greatest data burden, while use of intermittent video by both facilitator and learners can significantly lower data use, and thus costs. The choice of a platform also impacts teleprogramming, with creative options for use of lower cost platforms to reduce participant and training organization costs. These might include sharing educational content or video via chat groups and limiting conference to audio alone. In the context of COVID-19 where virtual meetings have become prevalent, it is critical that data burden is considered by program directors and funders. Looking forward, hybrid training that includes virtual and in-person training will likely become the norm in global health settings, but achieving this model will still require thoughtful consideration of data costs. Further, our findings are relevant to many other fields and advocate for evaluation of costs and data burden along with the growing use of teleprogramming in these settings.

**Author Summary:** The COVID-19 pandemic restricted travel and in-person gatherings. These restrictions also impacted access to important in-person training programs for healthcare workers, especially in areas where financial resources are limited. However, one positive impact of the pandemic has been improved access and experience with video conferencing tools (like Zoom) for many healthcare training programs. These video conferencing tools provide a way to complete essential training when in-person options may be limited. However, video-based training can have significant costs including internet data costs, mobile device costs, and healthcare worker professional time. Our study examined the data costs associated with video conferencing using several video conferencing applications across 5,554 mobile data plans in 228 countries. We found that costs are highest when trainers have their video on and learners participate with video on throughout the training. Intermittent video use by both trainers and learners can significantly lower costs. We also found that using lower-cost video-conferencing tools may help reduce costs. Additionally, there are training methods that can reduce costs including sharing educational content or video via chat groups and limiting conferences to audio only. Virtual training is a powerful and common tool in healthcare settings, but it is essential to consider costs, especially in areas with limited resources.

## Introduction

The COVID-19 pandemic has resulted in restrictions to travel and in-person gathering and reduced resources for training healthcare workers, particularly those in resource-scarce settings. At the same time, the increasing availability and accessibility of video conferencing platforms has led to their widespread use in local training events and national conferences. Initially, video conferencing could only be used by large organizations who could invest in devices, infrastructure and personnel to implement an on-premise solution that was made available to employees working on site. However, video conferencing systems are now widely available and accessible on a variety of devices including personal computers, smartphones, tablets, and even augmented reality headsets. Cloud videoconferencing solutions such as Zoom, Webex, Go-to-Meeting, Join.me, Telegram, Viber, WhatsApp, Team Viewer, and Microsoft Teams provide the essential video infrastructure as a fully supported hosted service. Many of these solutions also offer free services that allow video conferences between a limited number of participants for a finite duration, e.g. 45 minutes with participants using their own devices and paying for internet data costs. These solutions provide an option for delivering remote training when in-person options may be limited.

In healthcare settings, tele-education sessions may take the form of tele-mentoring guidance on the management of actual clinical cases or tele-simulation training [1-6]. Tele-simulation has been described in neonatal resuscitation training for healthcare workers in high and low resource settings [2, 7-8]. The Helping Babies Survive curriculum developed by the American Academy of Pediatrics is an evidence-based training curriculum that utilizes simulation principles to teach the steps of neonatal resuscitation and the routine care of small and sick newborns. The curriculum was designed for in-person facilitation, but also has been administered virtually to remote learners [8-9].

Some of the most visible demonstrations of tele-mentoring emerged from Project Extension for Community Healthcare Outcomes (ECHO), also known as Project ECHO, which utilizes tele-education to bridge knowledge gaps between specialists at academic health centers and primary care providers from remote areas [10]. The Project ECHO model expanded access to tele-mentoring through creating a virtual knowledge network on a range of topics related to care for patients. The first ECHO project linked university specialists with rural and prison-based clinicians to improve care for people with chronic hepatitis in New Mexico [11]. The concepts of tele-mentoring have also been applied to procedural training in surgical care [12-14] and are likely to have broad application to any field where skills training is required.

The cost considerations for digital training include internet data costs, mobile device costs and healthcare worker professional time. Many institutions are willing to support healthcare worker professional time to participate in training events and it is common for learners to own or have access to a mobile device. However, individuals in rural areas or low resource settings may not be able to access reliable broadband internet due to limited connectivity or high data costs. Despite the expansion of tele-simulation and tele-mentoring programs, little is known about the data costs to funding agencies and end-users. These expenses need to be taken into consideration as interest in delivery of remote training and mentoring grows in response to complex social and health demands as well as advancing technology.

## Methods

To assess data costs associated with digital training, two devices (an iPhone 7 and a Samsung Galaxy Note 8) were connected by video conference call for a 36-minute test period. All background apps and updates were disabled to accurately assess data use. Device A hosted the call and had video enabled throughout the test period (“continuous”) while Device B had video enabled for 2 minutes in 10 minutes intervals (“intermittent”). The test was performed for four platforms: Zoom, Webex, MS Teams and WhatsApp. The evaluation was performed with WIFI internet service and repeated using the dedicated sims of the two devices with a local mobile network provider (Globacom Nigeria). The bandwidth was monitored in real time using the MikroTik Userman hotspot monitor application to determine total data download and total data upload during the call for each device. Results were normalized to 60 minutes of use and the total time estimated for completing an essential newborn care provider course (approximated as 12 hours). Published country specific data on GB were accessed for analysis from: https://www.cable.co.uk/mobiles/worldwide-data-pricing/. These data were compiled from 5,554 mobile data plans in 228 countries (2020). The average cost of one gigabyte (1GB) was calculated and normalized to US dollars. Salary estimates were obtained from the Economic Research Institute (https://salaryexpert.com) and are based on salary survey data collected directly from employers and anonymous employees. Using the job description “registered nurse”, we conducted a search for the median hourly wage in each country and then converted the figures to 2021 USD.

## Results

### Data costs by country and region

The average cost of data as normalized to the price of 1 GB in USD varies greatly by country and region, ranging from 0.09 to 52.20 USD in 2020. Gross national income per capita also ranges nearly 500-fold from 280-117730 USD in 2019. Heat maps demonstrate that cost of data does not always parallel GNI (Fig 1). Cost analysis by region identified wide range with the highest (high) cost in Northern Africa and the lowest (low) cost in Asia and the CIS (USSR) (Table 1). The median of the average cost of 1GB across all regions was $3.42. The highest average cost of 1GB of data was in North America at $14/GB while the lowest average cost was in Northern Africa at $2/GB.

**Figure 1a.**
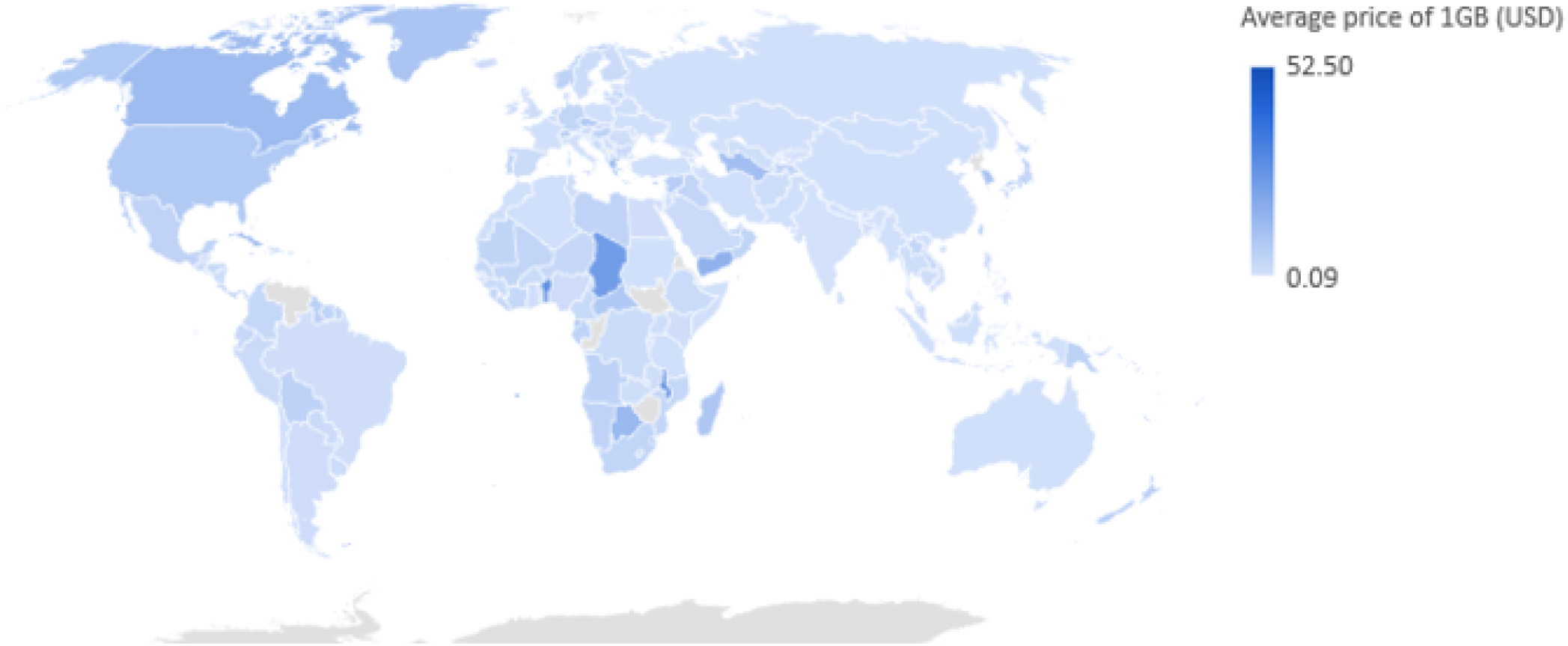
Average price of 1GB of data (USD)

**Figure 1b.**
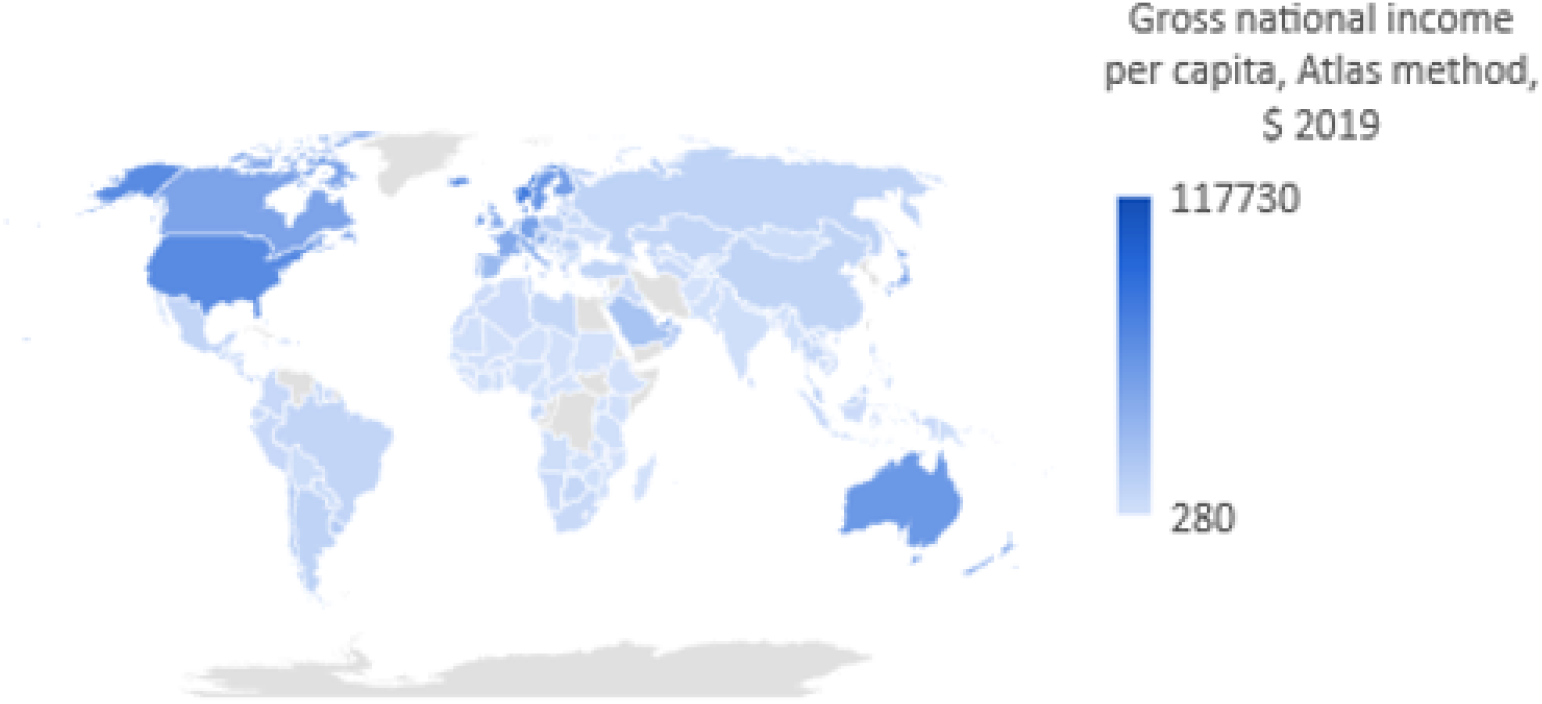
Gross national income per capita.

**Table 1.**
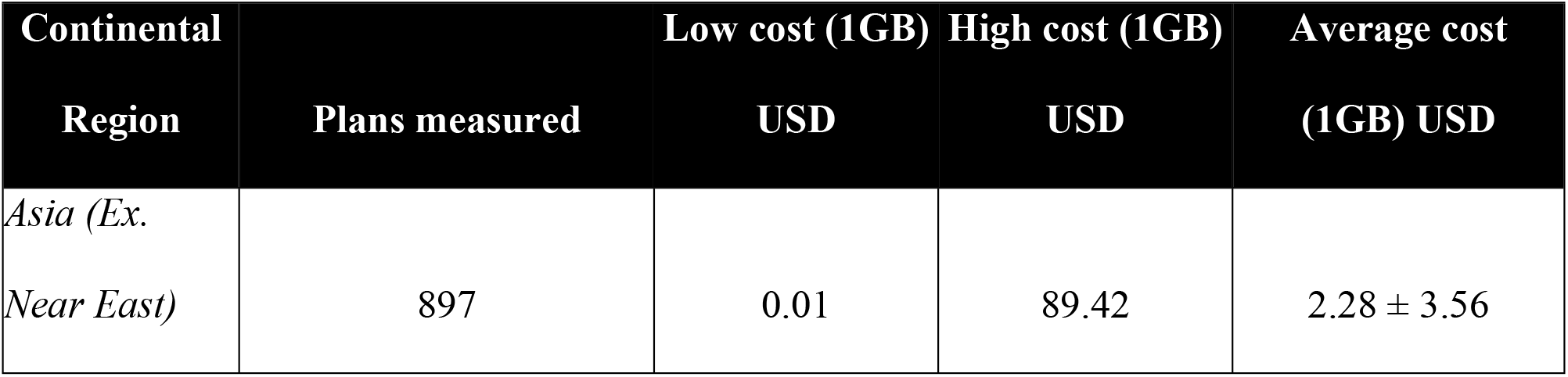
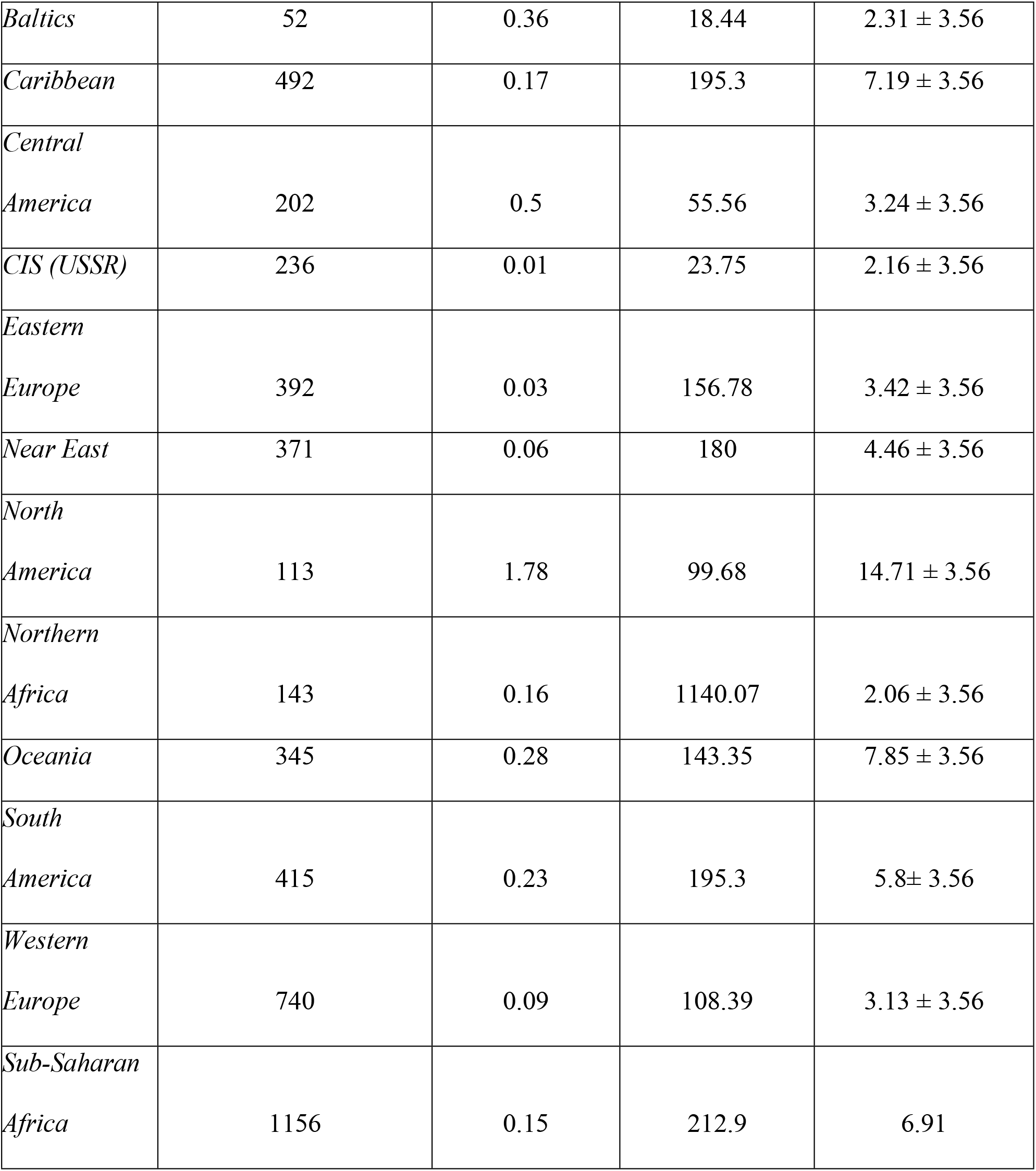
Regional Cost Ranges for 1GB of Data in U.S. Dollars.

### Data burden by connectivity and platform

To determine relative costs associated with use of variable platforms, the upload and download data use was determined modeling three typical training scenarios: 1) Continuous video for both facilitator and participant; 2) Continuous video for facilitator and intermittent video for participant; and 3) Intermittent video for both facilitator and participant. Representative platforms were assessed including Zoom, Webex (in high and low definition), Microsoft (MS) Teams and WhatsApp (Table 2).

**Table 2.**
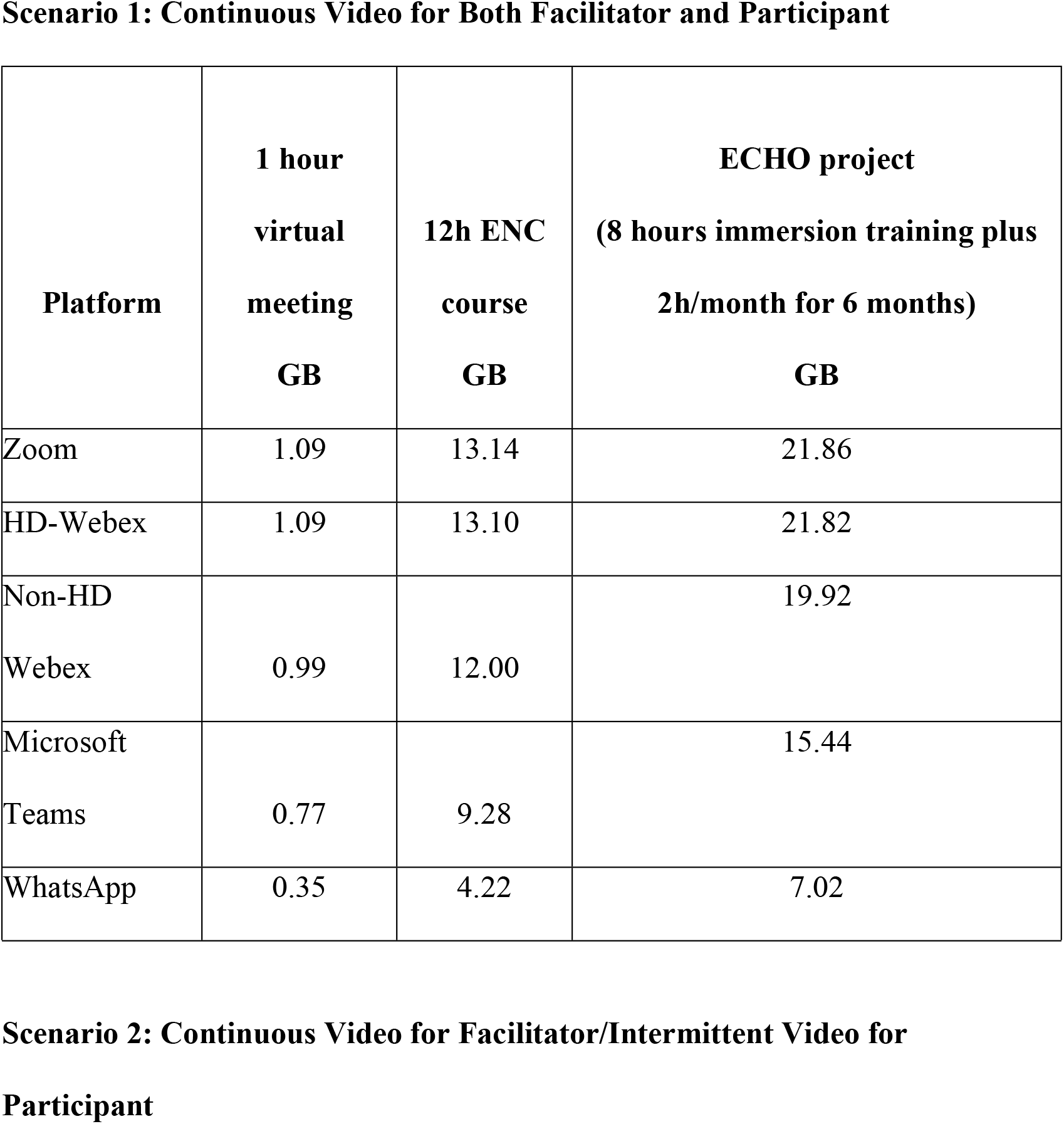
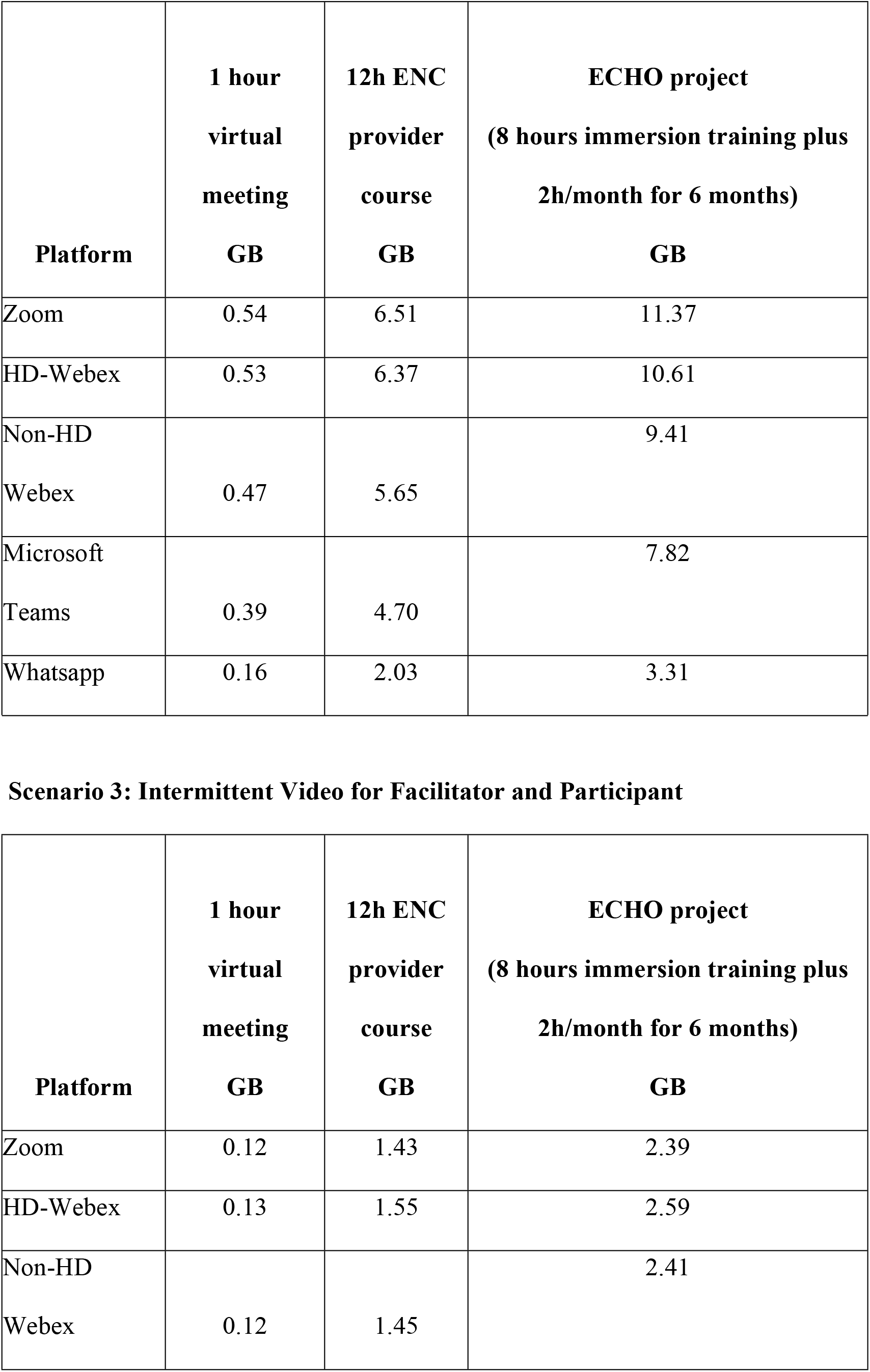
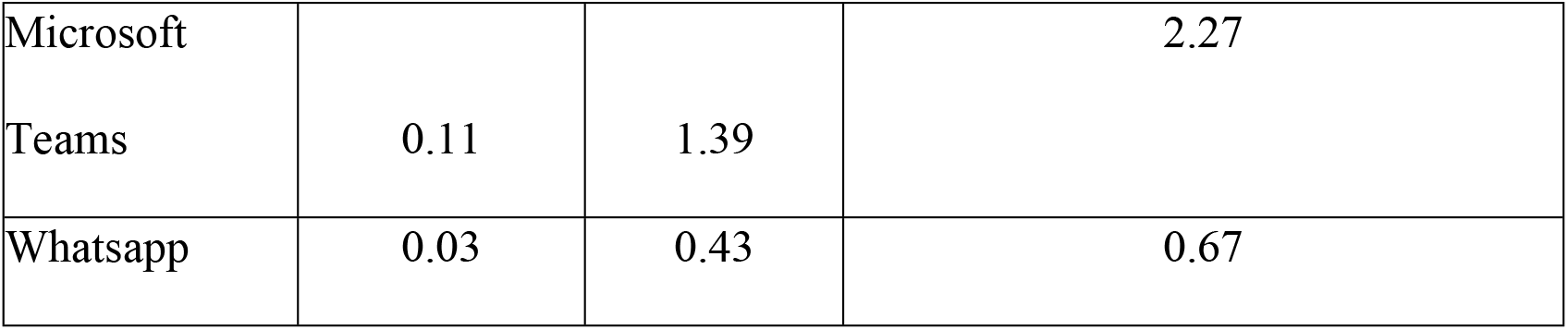
Comparison of Data Burden Associated with Variable Platforms and Continuous versus Intermittent Video Use.

### Earning wage and data equivalence by country

Average hourly wage for representative countries along with country specific GB costs were used to determine the relative number of work hours needed to cover the expense of participating in a 12-hour training session. These calculations identified a wide range, from greater than 4 hours in South Africa and Peru to less than 2 hours in Indonesia and Pakistan (Fig 2).

**Figure 2.**
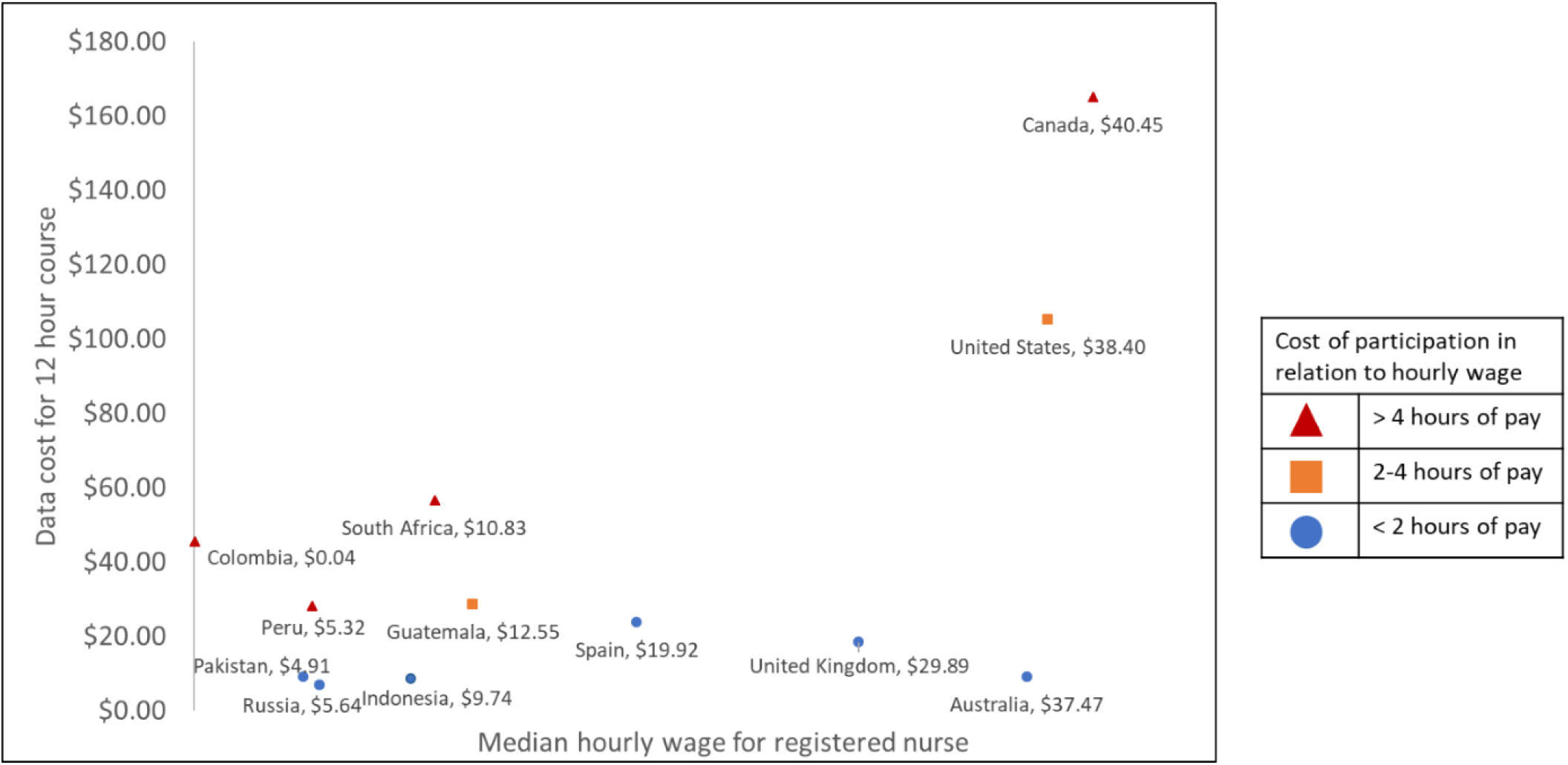
Estimated data cost for Zoom 12-hour course with continuous video by median hourly wage for a registered nurse.

## Discussion

Tele-education and tele-mentoring programs have been widely utilized and are of growing interest with need to provide safe training and education during the COVID-19 pandemic. With overburdened healthcare systems and restrictions on travel and in person gatherings, educators around the world have relied on online platforms to support international partnerships and to both, teach learners and provide clinical support through tele-mentoring [15-16]. Access to information and educational tools as well as challenges with internet access vary between urban and rural areas in high-income countries with even greater inequities in middle-to-low-income countries. Moreover, internet data costs vary significantly by country location in a way that do not mirror gross national income per capita exacerbating inequities and creating barriers to participation in tele-education and tele-mentoring. While institutions in high income countries often provide their staff with free or low-cost accounts to enable access to full-featured versions of the video conferencing platforms, many platforms also offer free, but technically limited versions. Access to joining calls is typically without a fee for attendees. Despite the “free” access, the cost of data packages may limit attendees’ ability to participate, particularly those on limited or “pay-as-you-go” internet or mobile phone plans that are prevalent in low resource settings.

Our data identifies that attendee expenses may be reduced by limiting the use of video for both the facilitator and the attendee. We found, not surprisingly, that the projected data cost is directly proportional to the use of video. However, in many cases, video is required to highlight key points or to observe performance of simulated procedures such as positive pressure ventilation during training in neonatal resuscitation or clinical findings with tele-mentoring. File compression or alternative sharing mechanisms may be helpful in the future. We found that costs could be limited significantly in scenarios where video was only used for brief periods of time by both the facilitator and the attendee. In a scenario where a dedicated interpreter might be required, the costs to the participant should be minimal if the interpreter does not share video. However, there will be a cost to the interpreter with use of video by both the facilitator and participant to provide adequate context to facilitate appropriate language interpretation.

Beyond limiting use of continuous video, alternative platforms which may have lower data burden due to differences in data encoding and transfer might be considered. We found that the use of Whatsapp [Meta] led to data costs that were one-third that of Zoom [Zoom Technologies]. However, there may be features that promote the use of one platform over another including security, restrictions on number of participants, user familiarity, or the availability of certain built-in features such as interpreters. Data conservation strategies may also be important in ensuring continued access when band-width is limited. In some countries, the penetrance of broadband is reasonable, but when a larger number of people are working from home, internet capacity may buckle.

In addition, the impact of the data costs must be considered in proportion to income of the attendees. With the significant variability in costs across both high- and low-income countries, it cannot be assumed that costs will be proportional to earned income in those countries. The cost to the end-user must be taken into consideration by programs and institutions seeking to utilize video conferencing broadly for education or clinical uses. A range of ECHO tele-mentoring projects have focused on pain management, rheumatology, geriatrics, hypertension, diabetes, behavioral health and palliative care [10, 17-18] A systematic review by Zhou et al., (2016) included 39 studies addressing 17 medical conditions. While evaluation was often limited, there were instances in which this approach changed provider behaviors, changed patient outcomes and was cost effective. [11] As these programs address the needs of more connected learners globally, it is important to consider the costs to both facilitators and learners and to design training programs that help to minimize these costs. Funders must consider the costs of data in their plans for implementation of new programs or in the transition from primarily in-person interactions to interventions that use technology to deliver education and support.

Communications advancements that have aided faster connections through increasing available bandwidth and reduced network loading have improved both the quality and capacity of video conferences. Thousands of networked callers can now be accommodated in multi-site meetings and conferences around the world. However, access to video conferencing platforms and reliable bandwidth to participate may vary. While tele-mentoring and tele-education might be assumed to represent distributed and equal access to education, application of these approaches in global health settings is complicated by structural variance in high- and low-income countries. Our study showed that there were significant differences between platforms and scenarios in terms of estimated data use which need to be taken into consideration where resources are limited.

Differences occur due to platform specific approaches to managing video and audio quality, bandwidth and security. Video standards or codecs determine the image quality and efficiency of video transmission within a program. A codec is a hardware or software-based process that compresses and decompresses large amounts of data used in voice over internet protocol video conferencing and streaming media. A codec takes data in one form, encodes it into another form and decodes it at the destination. Adaptive technologies deliver high quality video across networks with varying amounts of bandwidth. While some codecs send one video data stream for every resolution, frame rate and quality, others send just one stream that contains multiple layers of all the resolutions, frame rates and quality depending on what the devices and network that serve as the endpoints can support. This scalability supports meetings globally as each endpoint can select device and network-supported video resolution and quality without any additional encoding or decoding. The caller experiences an automatic degradation in the video quality when the network is busy while allowing the call to continue uninterrupted.

Platforms also differ with respect to security with many platforms employing encryption mechanisms to prevent unauthorized eavesdropping. However, higher levels of encryption can result in larger files and delays in transmission in audio and video, particularly with limited bandwidth, representing a greater data burden.

Our study had several limitations. We conducted the tests in one low resource setting; repeating the evaluation in multiple settings may have yielded additional results. We used two common mobile device brands (iPhone and Samsung), but use of additional device types may have helped to provide more generalizability. We intentionally focused on the internet data costs and did not consider attendee time, platform, or device costs. These all need to be thoughtfully included in the design and implementation of programs.

## Conclusion

The costs of participating in training programs that rely on video conferencing varies by location, mechanics of use, and the specific platform. We proposed practical solutions to limiting costs in low resource settings with the use of video conferencing calls. Scenarios in which facilitators have their video on and expect learners to participate with video on throughout result in the greatest data burden, while use of intermittent video by both facilitator and learners can significantly lower data use. The choice of a platform also impacts teleprogramming, with creative options for use of lower cost platforms to reduce costs. These might include sharing educational content or video via chat groups and limiting conference to audio alone. In the context of COVID-19 where virtual meetings have become prevalent, it is critical that data burden is adequately considered by program directors and funders. As hybrid virtual and in-person training become more common in global health settings, thoughtful consideration will need to be given to associated data costs. Our findings are relevant to many other fields and advocate for thoughtful evaluation of costs and data burden along with the growing use of teleprogramming in low resource settings.

## Data Availability

All data are included in the manuscript. Data available on request to Rachel Umoren.

